# Retrospective survey from Vascular Access Team Lombardy Net in COVID-19 era

**DOI:** 10.1101/2020.11.03.20222810

**Authors:** Gidaro Antonio, Vailati Davide, Gemma Marco, Lugli Francesca, Casella Francesco, Cogliati Chiara, Canelli Antonio, Nadia Cremonesi, Monolo Davide, Cordio Giuseppe, Frosi Chiara, Destefanis Riccardo, Rossi Anna, Alemanno Maria Chiara, Valenza Franco, Elli Stefano, Caldarini Andrea, Lucchini Alberto, Paglia Stefano, Baroni Monica, Giustivi Davide

**Affiliations:** Department of Biomedical and Clinical Sciences Luigi Sacco, University of Milan, Luigi Sacco Hospital, Milan, Italy; Intensive Care Unit ASST Melegnano Martesana; Intensive Care Unit Fatebenefratelli Hospital; UOC S.I.T.R.A. ASST OVEST MILANESE; Foundation Don Carlo Gnocchi Onlus Milano; Department of Oncology and Hemato-Oncology Fondazione IRCCS -Istituto Nazionale dei Tumor Milan; Intensive Care Unit ASST Monza; Emergency department ASST Lodi

**Keywords:** Venous Access Devices (VADs), COVID-19, Catheter Related Thrombosis (CRT), Catheter-Related Bloodstream Infection (CRBSI), Accidental Remove

## Abstract

**Background:** Venous Access Devices (VADs) are the most used device in COVID-19 patients.

**Objective:** Identify VADs implanted, catheter related thrombosis (CRT), catheter-related bloodstream infection (CRBSI), and accidental remove of VADs in both COVID-19 positive and COVID-19 free patients. Successive analysis was conducted comparing COVID-19 positive patients with COVID-19 free with inverse probability propensity score weights using simple regression to account for these two confounders (peripheral tip as central/peripheral and hospitalization as no/yes).

**Methods:** This multicenter, retrospective cohort study was conducted using data from 7 hospitals in Lombardy during the pandemic period from February 21^st^ to May 31^st^ 2020.

**Results:** 2206 VADs were evaluated, of which 1107 (50.2%) were inserted in COVID-19 patients. In COVID-19 cohort the first choice was Long Peripheral Cannula in 388 patients (35.1%) followed by Midline Catheter in 385 (34.8%). The number of “central tip” VADs inserted in COVID-free inpatients and COVID-19 positive were similar (307vs334). We recorded 42 (1.9%) CRT; 32 (79.2%) were observed in COVID-19 patients. 19 CRBSI were diagnosed; 15 (78.95%) were observed in COVID-19. Accidental removals were the more represented complication with 123 cases, 85 (69.1%) of them were in COVID-19. COVID-19 significantly predicted occurrence of CRT (OR = 2.00(1.85-5.03); P<0.001), CRSB (OR = 3.82(1.82-8.97); P<0.001), and Accidental Removal (OR = 2.39(1.80-3.20); P<0.001) in our propensity score weighted models.

**Conclusions:** CRT, CRBSI, and accidental removal are significantly more frequent in COVID-19 patients. Accidental removals are the principal complication, for this reason use of subcutaneously anchored securement is recommended for shorter period than usual.

## INTRODUCTION

Italy was the first western country where Coronavirus disease 2019 (COVID-19) spread, and the most affected region was Lombardy. Between February 21^st^, and May 31^st^ 2020, Lombardy counted 88968 COVID-19 patients the 38.2 percent^1^ of the whole country with a hospitalization rate of 35%. Venous Access Devices (VADs) are the most used device in COVID-19 patients both for nutrition and for therapies encountering the need of such complex and critical patients. During COVID-19 emergency, a group of experts from the Italian association GAVeCeLT (Gruppo Accessi Venosi Centrali a Lungo Termine) has formulated a few recommendations for the selection, insertion, and maintenance of the venous access devices, designed to protect the operator, to ensure the effectiveness of the maneuver, and to reduce the risk of complications.^2^ After the emergency, vascular access teams (VAT) of 7 hospitals in Lombardy (ASST FBF-Sacco; ASST Lodi; ASST Melegnano e Martesana; ASST Monza; ASST Ovest Milanese; Milan Istituto dei Tumori Hospital; Milan Fondazione Don Gnocchi Hospital) decided to perform an internal retrospective survey on all the VADs inserted in the period from February 21^st^ to May 31^st^ 2020. The main endpoint of the survey was to identify VADs type used during COVID-19 pandemic and catheter related complications such as catheter related thrombosis (CRT), catheter-related bloodstream infection (CRBSI) and accidental remove (AR) of VADs in COVID-19 positive and COVID-19 free patients; moreover, we compared the rates of these complications in COVID-19 positive and COVID-19 free patients.

## MATERIALS AND METHODS

### Study design and setting

In this multicenter, retrospective cohort study we collected and analyzed data regarding VADS’ insertion both in COVID-19 and non-COVID-19 patients in the period between February 21^st^ and May 31^st^. All COVID-19 patients were hospitalized, while non-COVID-19 could be either outpatients or inpatients. 7 Hospitals in Lombardy (ASST FBF-Sacco; ASST Lodi; ASST Melegnano e Martesana; ASST Monza; ASST Ovest Milanese; Milan Istituto dei Tumori Hospital; Milan Fondazione Don Gnocchi Hospital) were involved in this survey.

VADs insertion was performed following the CDC recommendations for vascular access in COVID-19 patients.^3^ The operator had to strictly adopt the “standard” maximal barrier precautions (hand hygiene, surgical mask, beret, sterile impermeable gown, sterile gloves, wide sterile drapes over the patient, and appropriate sterile cover for the ultrasound probe); furthermore, all the health workers that were physically present in the room wore either a N99 mask or a N95 mask and a surgical mask in addition to a face shield, during insertion procedure.

Ultrasound guidance was used for the insertion of all VADs.

VADs were divided into the following five categories based on length of catheter, insertion point and tip position, as indicated by Qin et al^4^: 1) Long Peripheral Cannula (LPC) (a.k.a. “Mini-Midline,” 6–15 cm), 2) Midline Catheter (MC, 15-25 cm), 3) Peripherally Inserted Central Catheter (PICC) (25-60 cm) 4) Femorally Inserted Central Catheter (FICC) 5) Centrally Inserted Central Catheter (CICC).

Catheter-related complications were collected and analyzed. Considered complications were: 1) CRT diagnosed with ultrasound^5^ or Computed Tomography (CT) with intravenous contrast 2) CRBSI diagnosed with blood culture performed by the catheter that showed microbial growth at least two hours earlier than growth detected in blood collected simultaneously from a peripheral vein.^6^ 3) accidental removals when patients or healthcare personnel accidentally pulled out the catheter.

### Ethical approval

The study (“REGISTRO DELLE INFEZIONI SOSPETTE E ACCERTATE COVID-19/Studio Sacco COVID-19)” was approved by the local ethical committee Milan Area 1 in ASST Fatebenefratelli Sacco, University Hospital Luigi Sacco with the registration number 2020/16088.

### Statistical Analysis

Categorical variables are reported as number (percentage). Continuous variables are reported as mean±SD[median(IQR)].

Since the majority of COVID-19 patients received “peripheral tip” catheters (773 (69.8%) patients) and all of them were hospitalized, we calculated inverse probability propensity score weights using simple regression to account for these two confounders (peripheral tip as central/peripheral and hospitalization as no/yes).

Three logistic models were built to assess the predictivity of COVID-19 disease respectively towards CRT, CRBSI, and Accidental Removal using the calculated weights. The Average Treatment Effect (ATE) was estimated.

Odds Ratios are reported together with their 95% CIs.

The statistical analysis was performed with R version 4.0.0 (2020-04-24) -- “Arbor Day” Copyright (C) 2020 The R Foundation for Statistical Computing Platform: x86_64-w64-mingw32/x64 (64-bit) - Vienna, Austria. URL https://www.R-project.org/.

## RESULTS

2206 VADs were evaluated (Table 1), of which 1107 (50.2%) were inserted in COVID-19 patients. The most used VADs in COVID-19 free cohort were PICC which were inserted in 480 patients (43.9%) followed by MCs in 465 patients (42.3%). In COVID-19 cohort the first choice was LPC in 388 patients (35.1%) followed by MCs in 385 patients (34.8%).

**TABLE 1:** Total number of devices with complication; COVID-19 free device with complication; COVID-19 device with complication. CICC= Centrally Inserted Central Catheter (Internal Jugular Vein, Axillary Vein, Subclavian Vein, Anonymous Vein) FICC= Femorally Inserted Central Catheter PICC= Peripherally Inserted Central Catheter CRT= catheter related thrombosis CRBSI=catheter-related bloodstream infection

| TOTAL DEVICE | N°(%) | CRT (%) | CRBSI (%) | ACCIDENTAL REMOVALS |
| --- | --- | --- | --- | --- |
| CICC | 159<br>(159/2206 7,2%) | 2<br>(2/159 1,3%) | 3<br>(3/159 1,9%) | 10<br>(10/159 6,3%) |
| FICC | 85<br>(85/2206 3,9%) | 3<br>(3/85 3,5%) | 4<br>(4/85 4,7%) | 10<br>(10/85 11,8%) |
| PICC | 608<br>(608/2206 27,6%) | 8<br>(8/608 1,3%) | 2<br>(2/608 0,3%) | 20<br>(20/608 3,3%) |
| Midline | 850<br>(848/2206 38,5%) | 18<br>(18/848 2,2%) | 8<br>(8/848 0,9%) | 36<br>(36/848 4,2%) |
| Long Periph.<br>Cannula | 504<br>(500/2206 22,7%) | 11<br>(11/500 2,2%) | 2<br>(2/500 0,4%) | 47<br>(47/500 9,5%) |
| tot | 2206 | 42<br>(42/2206 1,9%) | 19<br>(19/2206 0,9%) | 123<br>(123/2206 5,6%) |
| COVID-19 FREE DEVICE | N°(%) | CRT (%) | CRBSI (%) | ACCIDENTAL REMOVALS |
| CICC | 17<br>(17/1099 1,5%) | 0 | 0 | 1<br>(1/17 5,9%) |
| FICC | 21<br>(21/1099 1,9%) | 1<br>(1/21 4,8%) | 0 | 3<br>(3/21 14,3%) |
| PICC<br>Inpatients | 269<br>(269/1099 24,5%) | 1<br>(1/269 0,4%) | 0 | 5<br>(5/269 1,9%) |
| PICC<br>Outpatients | 211<br>(211/1099 19,2%) | 1<br>(1/211 0,5%) | 1<br>(1/211 0,5%) | 10<br>(10/211 4,7%) |
| Midline<br>Inpatients | 452<br>(452/1099 41,1%) | 4<br>(4/452 0,9%) | 3<br>(3/452 0,7%) | 13<br>(13/452 2,9%) |
| Midline<br>Outpatients | 13<br>(13/1099 1,2%) | 0 | 0 | 0 |
| Long Periph.<br>Cannula | 116<br>(112/1099 10,6%) | 3<br>(3/112 2,7%) | 0 | 6<br>(6/112 5,4%) |
| tot | 1099 | 10<br>(10/1099 0,9%) | 4<br>(4/1099 0,4%) | 38<br>(38/1099 3,5%) |
| COVID-19 DEVICE | N°(%) | CRT (%) | CRBSI (%) | ACCIDENTAL REMOVALS |
| CICC | 142<br>(142/1107 12.8%) | 2<br>(2/142 1.4%) | 3<br>(3/142 2,13%) | 9<br>(9/142 6.3%) |
| FICC | 64<br>(64/1107 5.8%) | 2<br>(2/64 3.1%) | 4<br>(4/64 6,25%) | 7<br>(7/64 10.9%) |
| PICC | 128<br>(128/1107 11.5%) | 6<br>(6/128 4.7%) | 1<br>(1/128 0,78%) | 5<br>(5/128 3.9%) |
| Midline | 385<br>(385/1107 34.8%) | 14<br>(14/385 3.6%) | 5<br>(5/385 1,3%) | 23<br>(23/385 6%) |
| Long Periph.<br>Cannula | 388<br>(388/1107 35.1%) | 8<br>(8/388 2.1%) | 2<br>(2/388 0,52%) | 41<br>(41/388 10.6%) |
| tot | 1107 | 32<br>(32/1107 2.9%) | 15<br>(15/1107 1.4%) | 85<br>(85/1107 7.7%) |

The majority of COVID-19 patients received “peripheral tip” catheters (773 (69.8%) patients) and all of them were hospitalized,

COVID-19 free patients implanted 581 (52.9%) “peripheral tip” catheters. It has to be emphasized that in COVID-19 free cohort 224 (20.4%) VADs were inserted in outpatients: 211 PICC (94.2%)with “central tip”; 13 MCs “peripheral tip”. When considering the “central tip” venous accesses, inserted in COVID-free inpatients the number was similar as compared to COVID-19 positive patients (respectively 307 vs 334).

In the observation period we recorded 42 (1.9%) CRT; 32 (79.2%) were observed in COVID-19 patients. Only one COVID-19 patient develop pulmonary embolism (Table 2).

**TABLE 2.** Description of VADs complication

|  | Number of complication | Male /Female % | Median Age | Median day insertion/diagnosis | pulmonary embolism | CRBSI patogen | New VAD after Accidental remove | Mortality after complication |
| --- | --- | --- | --- | --- | --- | --- | --- | --- |
| CRT COVID+ | 32 | 50%/50% | 69.7±11.2 | 8.7±5.7 | 1 |  |  | 31.3% |
| CRT COVID FREE | 10 | 50%/50% | 67.1±12.7 | 15.6±18.1 | 0 |  |  | 20.0% |
|  | p <0.001 |  |  |  |  |  |  |  |
| CRBSI COVID+ | 15 | 40%/60% | 63.2±19.7 | 17.2±8.3 |  | GRAM + 11/ GRAM - 3/ Fungal 1 |  | 20% |
| CRBSI COVID FREE | 4 | 40%/60% | 62 ± 15.6 | 39.7± 50.5 |  | GRAM + 2/ GRAM - 1/ Fungal 1 |  | 0% |
|  | p <0.001 |  |  |  |  |  |  |  |
| Accidental Remove COVID+ | 85 | 55%/45% | 77.5±12.5 | 7±6.3 |  |  | 35% | 30% |
| Accidental Remove COVID FREE | 38 | 37%/63% | 70.9±15.8 | 16.8±17.9 |  |  | 44% | 12% |
|  | p <0.001 |  |  |  |  |  |  |  |

19 CRBSI were diagnosed. 15 were observed in the COVID-19 group (78.95%). The majority of CRBSI were due to GRAM positive bacteria. (Table 2)

Accidental removals were the most represented complication with 123 registered cases, 85 of them (69.1%) were in COVID-19 (Table 2).

COVID-19 significantly predicted the occurrence of CRT (OR = 2.00(1.85-5.03); P<0.001), CRSB (OR = 3.82(1.82-8.97); P<0.001), and Accidental Removal (OR = 2.39(1.80-3.20); P<0.001) in our propensity score weighted models.

## DISCUSSION

To the best of our knowledge this is the first survey on VADs in COVID-19 patients.

VAT staff during the pandemic split their time both for COVID-19 and COVID-19 free patients with a good balance as explained to the same number of VADs in the two groups. It is important to not underestimate the increase of workload and the work-related stress for healthcare staff in this pandemic period. Moreover, the psychological stress was relevant considering the reduction of human resources. However, the members of VATs worked at their best to assist all patients, adopting personal protective equipment, and breaking down days of absence from vascular access teams during this emergency period.

In our study VADs type used for COVID-19 patients were in agreement with GAVeCeLT^2^ indication recommending MCs and LCP as first choice in this setting. Indeed, due to their longer dwell time, MC and LCP can reduce the number of peripheral venous insertions required thus saving resources and reducing infective risks for the operator; these are definite advantages in COVID-19 pandemic.

When considering the central tip venous accesses in inpatients, the number was similar comparing COVID-19 free and COVID-19 positive patients (respectively 307 vs 334). Nevertheless, we observed a significantly increased use of CICC and FICC in COVID-19 patients as compared to COVID-19 free patients. The increase number of inserted FICC is probably justified by the minimal interference with respiratory management in patients on CPAP/NIV and by the reduced risk of operator contamination by the patient’s oral, nasal, and tracheal secretions during insertion. On the contrary we observed a reduced number of inserted PICC in

COVID-19 cohort which can be partially explained by difficult catheterization due to upper limb edema in patients on CPAP and the perceived increased risk of upper limb vein thrombosis. Furthermore, PICC are usually single lumen, only a few are double, while CICC and FICC normally have more than one lumen because they are used in intensive care units where multiple infusion lines are needed.

In this study population, CRT rates in non-COVID-19 patients were lower than those reported in literature,^7,8^ and COVID-19 patients had a statistical higher rate of CRT as compared with COVID-19 free. This finding is consistent with recent studies, indicating that venous thrombotic risk of patients with COVID-19 is higher than normal, despite the presence of prophylactic heparin therapy.^9^ The increased thrombotic risk of VADs in COVID-19 patients is mainly driven by the higher CRT incidence in peripherally inserted devices such as PICCs and MCs. One of the possible explanations may be the increased upper limb venous stasis due to underarm fastening straps of helmet used during ventilation with continuous positive airway pressure (CPAP).^10^ As a consequence, the use of alternative strategies to reduce CRT such as intermediate (50-70 units/kg/12 h) or therapeutic dose of low-molecular weight heparin (100units/kg/12h or 150units/kg/24h) should be considered in all COVID-19 patients with central or peripheral inserted VADS with high thrombotic risk.^2^

CRBSI in our study was higher for COVID-19 than non-COVID-19 patients but overall lower incidence than what reported in literature ^11,12^. This could be due to the large use of parenteral nutrition (PN) through VADs during pandemic. PN was found to be an independent risk factor for catheter infection^13^. In a recent paper about COVID-19 CRBSI three out of six patients have candida, two have GRAM positive bloodstream infection^14^. Fungal and gram positive infections are a particular concern in patients receiving a high concentration of glucose in intravenous hyperalimentation occurring more frequently in immunosuppressed patients and in patients who have received multiple antibacterial antibiotics such as many COVID-19 patients (Azithromycin, Tocilizumab and Steroids were the standard of care during the first month of pandemic). The preference of PN over enteral nutrition (EN), which may certainly be debatable, is probably due to the use of non-invasive ventilation through Helmet interface or endotracheal intubation. Another important reason is the fear of the operators to become infected with COVID-19, that could translate in poor care of the VADs to reduce the contact time with COVID-19 patients.^15^

Accidental removals were the most frequent complication in the study population: incidence of cases is higher in COVID-19 patients even when compared with literature data.^16^ COVID-19 patients are often at risk for dislodgment particularly during the maneuvers of pronation-supination.^17^ This finding is also influenced by psychiatric complications of COVID 19 disease, particularly delirium, that appears to be common in critically ill patients. In one series of 58 patients with COVID-19-related acute respiratory distress syndrome (ARDS), delirium/encephalopathy was present in approximately two-thirds of patients.^18^ Delirium is a recognized risk factor for vascular access removal.^19^ The insertion of medium and long-term indwell times VADs and the practice of using subcutaneously anchored securement for medium and long-term indwell times VADS while reserving adhesive devices if catheter indwell times is lower than 7 days may reduce the incidence of accidental removals^20^.

Our study has several limitations. First of all, we compared the rate of VADs complications in two populations (COVID 19 positive and COVID free patients) which are probably different in terms of risk factors for CRT, CRBSI and AR. We reduced the effects of selection biases by using inverse probability propensity score weights to account for two confounders which significantly differed among the two populations (peripheral tip as central/peripheral and hospitalization as no/yes). However other potential demographic and clinical confounders should probably be taken into account when comparing such different populations.

In our study we did not collect demographic and clinical data that could potentially represent risk factors for VADs complications; this is definitely another limitation of this investigation.

Finally, our study is retrospective: the used VADs were chosen by the clinician for each patient and subsequently changed on the basis of clinical experience and data emerging from literature.

## CONCLUSION

Issues related to venous thrombotic risk, type of ventilatory support and the potential operators’ contamination can influence the choice of the most appropriate VAD in patients with COVID 19 patients.

CRT, CRBSI, and accidental removal are significantly more frequent in COVID-19 patients. The expertise and the experience of VATs team can potentially reduce the incidence of complications with a wisely choice of VADs balancing patients characteristic, drug infused, disease course, thanks to the effective use of best practice guidelines and the correct use of the instrumentation. Accidental removals are the principal complication; for this reason, the use of subcutaneously anchored securement is recommended for shorter period than usual.

The Authors declare no conflicts of interest, no financial support.

The data supporting the findings of this study are available from the corresponding author upon reasonable request.

## References

1) Italian Ministry of Health, Italian civil protection site http://opendatadpc.maps.arcgis.com/apps/opsdashboard/index.html#/b0c68bce2cce478eaac82fe38d4138b1

2) Pittiruti M, Pinelli F; GAVeCeLT Working Group for Vascular Access in COVID-19. Recommendations for the use of vascular access in the COVID-19 patients: an Italian perspective. Crit Care. 2020;24(1):269. Published 2020 May 28. doi:10.1186/s13054-020-02997-1

3) CDC. Updated protocol on airborne precautions, 2020, https://www.cdc.gov/coronavirus/2019-ncov/hcp/clinicalguidance-management-patients.html

4) Qin KR, Pittiruti M, Nataraja RM, Pacilli M. Long peripheral catheters and midline catheters: Insights from a survey of vascular access specialists. J Vasc Access. 2020 Oct 20:1129729820966226. doi: 10.1177/1129729820966226. Epub ahead of print. PMID: 33078685.

5) Sartori M, Migliaccio L, Favaretto E, et al. Whole-Arm Ultrasound to Rule Out Suspected Upper-Extremity Deep Venous Thrombosis in Outpatients. JAMA Intern Med. 2015;175(7):1226–1227. doi:10.1001/jamainternmed.2015.1683

6) National Healthcare Safety Network. Bloodstream Infection Event (Central Line-Associated Bloodstream Infection and Non-Central Line-Associated Bloodstream Infection). http://www.cdc.gov/nhsn/PDFs/pscManual/4PSC_CLABScurrent.pdf (xAccessed on November 16, 2015).

7) Chopra V, Kaatz S, Swaminathan L, et al. Variation in use and outcomes related to midline catheters: results from a multicentre pilot study. BMJ Qual Saf. 2019;28(9):714–720. doi:10.1136/bmjqs-2018-008554

8) Balsorano P, Virgili G, Villa G, et al. Peripherally inserted central catheter-related thrombosis rate in modern vascular access era-when insertion technique matters: A systematic review and meta-analysis. J Vasc Access. 2020;21(1):45–54. doi:10.1177/1129729819852203

9) Bilaloglu S, Aphinyanaphongs Y, Jones S, Iturrate E, Hochman J, Berger JS. Thrombosis in Hospitalized Patients With COVID-19 in a New York City Health System [published online ahead of print, 2020 Jul 20]. JAMA. 2020;e2013372. doi:10.1001/jama.2020.13372

10) Vailati D, Fusco T, Canelli A, Chiariello C, Zerla P. Axillary vein thrombosis in COVID positive patient with midline and continuous positive airway pressure Helmet [published online ahead of print, 2020 Jul 15]. J Vasc Access. 2020;1129729820943424. doi:10.1177/1129729820943424

11) European Centre for Disease Prevention and Control. Surveillance report: point prevalence survey of healthcare associated infections and antimicrobial use in European acute care hospitals. Stockholm: ECDC, 2013.; Available from: http://ecdc.europa.eu/en/publications/Publications/healthcare-associated-infections-antimicrobial-use-PPS.pdf.

12) Blanco-Mavillard I, Rodríguez-Calero MÁ, de Pedro-Gómez J, Parra-García G, Fernández-Fernández I, Castro-Sánchez E. Incidence of peripheral intravenous catheter failure among inpatients: variability between microbiological data and clinical signs and symptoms. Antimicrob Resist Infect Control. 2019;8:124. Published 2019 Jul 22. doi:10.1186/s13756-019-0581-8

13) Reitzel RA, Rosenblatt J, Chaftari AM, Raad II. Epidemiology of Infectious and Noninfectious Catheter Complications in Patients Receiving Home Parenteral Nutrition: A Systematic Review and Meta-Analysis. JPEN J Parenter Enteral Nutr. 2019;43(7):832–851. doi:10.1002/jpen.1609

14) Hughes S, Troise O, Donaldson H, Mughal N, Moore LS. Bacterial and fungal coinfection among hospitalised patients with COVID-19: A retrospective cohort study in a UK secondary care setting [published online ahead of print, 2020 Jun 27]. Clin Microbiol Infect. 2020;S1198-743X(20)30369-4. doi:10.1016/j.cmi.2020.06.025

15) Kumar J, Katto MS, Siddiqui AA, Sahito B, Ahmed B, Jamil M, Ali M. Predictive Factors Associated With Fear Faced by Healthcare Workers During COVID-19 Pandemic: A Questionnaire-Based Study. Cureus. 2020 Aug 14;12(8):e9741. doi: 10.7759/cureus.9741. PMID: 32944456; PMCID: PMC7489766.

16) Chopra V, Kaatz S, Swaminathan L, et al. Variation in use and outcomes related to midline catheters: results from a multicentre pilot study. BMJ Qual Saf. 2019;28(9):714–720. doi:10.1136/bmjqs-2018-008554

17) Elharrar X, Trigui Y, Dols AM, et al. Use of Prone Positioning in Nonintubated Patients With COVID-19 and Hypoxemic Acute Respiratory Failure JAMA. 2020;323(22):2336–2338. doi:10.1001/jama.2020.8255

18) Helms J, Kremer S, Merdji H, et al. Neurologic Features in Severe SARS-CoV-2 Infection. N Engl J Med. 2020;382(23):2268–2270. doi:10.1056/NEJMc2008597

19) Sundararajan K, Wills S, Chacko B, Kanabar G, O’Connor S, Deane AM. Impact of delirium and suture-less securement on accidental vascular catheter removal in the ICU. Anaesth Intensive Care. 2014;42(4):473–479. doi:10.1177/0310057X1404200408

20) Macmillan T, Pennington M, Summers JA, et al. SecurAcath for Securing Peripherally Inserted Central Catheters: A NICE Medical Technology Guidance. Appl Health Econ Health Policy. 2018;16(6):779–791. doi:10.1007/s40258-018-0427-1

